# Genetic decoding reveals druggable biology implicitly learned by a medical-history foundation model

**DOI:** 10.64898/2026.07.28.26359117

**Authors:** Wenhuan Zeng, Leonhard Kohleick, Carl Beuchel, Martijn Zoodsma, Mine Koprulu, Julia Carrasco Zanini, Benjamin Wild, Claudia Langenberg, Maik Pietzner

**Affiliations:** Health Data Modelling, Berlin Institute of Health at Charité – Universitätsmedizin Berlin, Berlin, Germany; Computational Medicine, Berlin Institute of Health at Charité - Universitätsmedizin Berlin, Berlin, Germany; Precision Healthcare University Research Institute, Queen Mary University of London, London, United Kingdom; Digital Health, Berlin Institute of Health at Charité – Universitätsmedizin Berlin, Berlin, Germany; Institut für Medizinische Informatik, Charité – Universitätsmedizin Berlin, Berlin, Germany

## Abstract

Foundation models trained on electronic healthcare records (EHRs) have gained traction with the aim to transform personalised medicine. However, their interpretability is bound to redescribing the records the models were trained on, missing implicitly learned concepts and biases. Here, we show that human genetics provides an orthogonal layer to surface implicitly learned biological concepts and otherwise hidden risk factors. Re-implementing the generative transformer Delphi-2M in >500,000 UK Biobank participants, we performed genome-wide association testing on its 120 learned embeddings and identified 434 genome-wide-significant signals across 151 independent loci and 98 embeddings, revealing a heritable structure that feature-attribution methods cannot recover. Effector-gene mapping implicated cholesterol metabolism and an IL-1-family epithelial-alarmin pathway, supported by strong (>50-fold) enrichment for variants previously associated with blood lipids, body-mass index, and asthma. Loci recovered the targets of essentially all approved lipid-lowering and severe-asthma therapies, and another twelve drugs not obvious from genetic results based on single ICD-10 GWAS. Yet, embeddings poorly explained variation in pleiotropic risk factors, while still retaining most of their predictive value. Substantial improvements in predictive performance were hence confined to a minority of common diseases by adding specific diagnostic or organ-derived markers. Our findings suggest that human genetics might be most powerful as an orthogonal explanatory or regularising layer to train the next generation of EHR-based foundation models that likely benefit most from the addition of targeted biomarkers to advance personalised medicine.

## INTRODUCTION

Foundation models trained on longitudinal electronic health records (EHRs) can predict the onset of hundreds of diseases from a person’s prior disease trajectory, learning lifetime morbidity rather than isolated diagnoses^1^. We^2^ and others^3–5^ have shown that medical history captured in the course of healthcare predicted disease onset across specialties and transferred across healthcare systems. The generative transformer Delphi-2M is among the most recent, providing calibrated forecasts for more than 1,000 diseases across the life course^6^. Deployment in clinical settings, however, requires a detailed understanding of what context those models have learned and utilize to predict, explicitly based on input features, so-called tokens, but also implicitly based on hidden structures in the data (**Fig. 1a**). The latter is of utmost interest, since it may reveal concepts of risk missed by traditional approaches but also hint at biases that propagate inequalities in healthcare.

**Figure 1.**
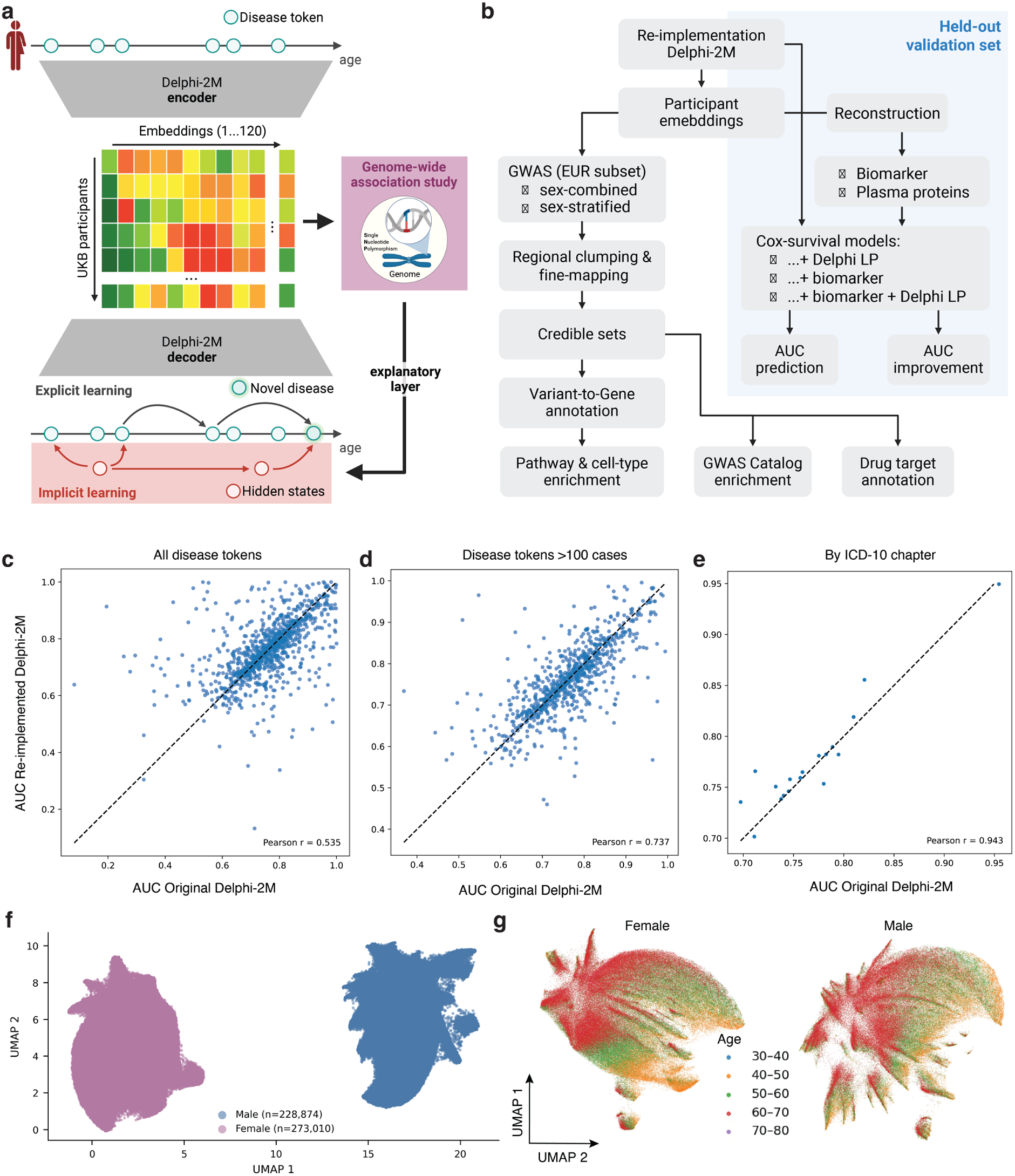
Conceptual framework and study design to explain Delphi-2M predictions. **a** Simplified concept of how Delphi-2M transforms a sequence of diagnoses into numerical representations that enable the prediction of the onset of a novel disease in a participant. Genome-wide associations studies (GWAS) are placed as an explanatory layer to understand implicitly learned concepts not recoverable from input tokens, that is diseases. Individual-level embeddings were generated by mean-pooling the final-layer hidden states across all non-padding tokens, yielding a 120-dimensional embedding vector describing the entire health trajectory of each participant. **b** Overview of the performed analysis. LP = linear predictor; AUC = area under the receiver operating curve. **c-e** Scatterplots opposing the AUC reported in the original paper (x-axis) and base on reimplementation (y-axis). **f-g** UMAP projections of the derived participant embeddings coloured by sex (**f**) and stratified by sex (**g**) and coloured by age groups. **a** and **b** were created with BioRender.com

Current explainability methods are largely confined to feature-attribution techniques such as SHAP (SHapley Additive exPlanations), which allocate a single prediction back onto the input tokens the model was trained on^7^. While informative, attribution techniques are fundamentally restricted since they can only redescribe relationships among EHRs the model was trained on and cannot reveal the latent structure a model may have implicitly learned that represents risk factors and putative biases (**Fig. 1a**). For example, we recently showed that associating risk predictions based on a retinal image foundation model with participant’s genetic information revealed strongly predicted features, such as eye colour, not inferable from feature attribution techniques based on the supplied images^8^. However, genome-wide association testing has been so far almost exclusively used as discovery rather than explanatory tool^9–11^. Most recently, a Bayesian framework performed genome-wide association testing on latent disease signatures learned jointly from longitudinal EHRs and polygenic scores, recovering pleiotropic cardiometabolic loci not evident in single-trait analyses^12^. However, genetic information was itself part of the model’s input and the aim was discovery and prediction. In contrast, utilizing genetic data to explain rather than to augment prediction models is supported by the confined predictive value even when cumulating effects across the entire genome for most common diseases^13^ and its high recovery rate of successful drug targets^14^. We here therefore investigated whether genetic information, fixed at conception, least affected by reverse causation, and never supplied to the model, can help surface implicitly learned concepts and biases to derive an explainable artificial intelligence framework agnostic to the model architecture and input data modalities.

To this end, we re-implemented Delphi-2M in >500,000 UK Biobank participants and derived 120 learned patient-level embeddings as quantitative phenotypes for genome-wide association testing (**Fig. 1b**). We report significant heritability that recovered implicitly learned domains of risk, including a druggable domain of lifetime morbidity, but also evidence of biases. We finally show that such models are likely to benefit most from selected disease-specific biomarker to improve predictive performance^15^. Our results support the use of human genetics as an orthogonal layer to explain, rather than necessarily augment, EHR-derived foundation models to improve our understanding in how such models come to decisions that may inform healthcare practice.

## RESULTS

### Patient-level embeddings cluster by sex and age but not clinical specialty

We first re-implemented the Delphi-2M on our UK Biobank (application no. 44448) release since model weights from the original implementation were not available at the time of the study. Briefly, we adopted the training procedure described by the authors using the same ICD-10 coded first occurrence diagnostic data. We benchmarked successful implementation by demonstrating 1) similar clustering of disease tokens space by ICD-10 category when projecting them using Uniform Manifold Approximation and Projection (UMAP)^16^ (**Supplementary Fig. 1**), and 2) a consistent tendency in model performance across all diseases (Pearson correlation of AUC values = 0.53; **Fig. 1c-e**), with those diseases already predicted poorly and those with low cases numbers expected to vary the most (Pearson correlation of AUC values improved to 0.74 upon exclusion of rare outcomes) (**Fig. 1c-d**). We note that the average performance across ICD-10 chapters were comparable (mean AUC (origin) = 0.757; mean AUC (replication) = 0.762). We next derived 120 participant-level embeddings, the latent numeric space generated by the model from text input to make predictions, by using the model as an encoder (**Fig. 1a**). Embeddings were not completely independent of each other, and we observed ten clusters likely representing similar domains of participants’ health (**Supplementary Fig. 2**). We note, that the derived individual-wise embeddings were not used or displayed in the original publication to understand the context learned by Delphi-2M.

To understand what biological information such patient-derived embeddings hold, we first projected them into a two-dimensional space using UMAP. We observed a complete separation of the sexes in this latent space (**Fig. 1f**), and further a strong age gradient within each sex (**Fig. 1g**), but a lack of clear separation by diagnoses (**Supplementary Fig. 3**). Since the relevance of input tokens, that is disease diagnoses, has already been presented in great detail in the original publication^6^, we directly proceeded to genome-wide association testing.

### Genetic architecture and causal variant prioritization of patient-level embeddings

We first tested for an association between 14,081,025 high-quality genetic variants (minor allele frequency (MAF) > 0.5%) with each of the 120 derived embeddings among UKB participants of British European ancestry (n=441,189). We observed significant (p<0.05/120) narrow sense SNP-based heritability estimates on the observed quantitative scale (h^2^) for all embeddings with a median of 1.9% (IQR:1.4%-2.4%), reaching values up to 6.35% for embedding36 (**Supplementary Tab. 2**). Values that were largely in line with estimates for single disease that served as input on the same scale and even marginally higher compared to recently reported signature loadings^12^. We note, however, that a direct comparison with liability-scale estimates for the binary diseases that served as model input is only approximate, since the two are expressed on different scales. We therefore treat the correspondence as qualitative support for the biological validity of the embeddings to understand the biological context of lifetime multimorbidity learned by the model rather than as a quantitative equivalence.

We discovered a total of 434 significant (p<5×10^−8^) independent genetic variant – embedding associations (**Fig. 2 and Supplementary Tab. 3**) based on a combination of regional clumping (±500kb) and Bayesian fine-mapping^17^, treating the extended MHC region separately. Out of those, almost all (n=419) also passed a more stringent significance threshold (p<4.2×10^−10^) controlling for the number of tested embeddings, indicating general robustness of our findings. A total of 98 embeddings showed at least one associated lead credible set variant (median: 2; IQR: 1 - 6). Five embeddings associated with 15 or more distinct variants (embeddings in decreasing order of associated variants: 92, 37, 64, 4, and 29) indicating a polygenic background (**Fig. 2**). Association results followed the typical negative, log-inverse relationship between MAF and absolute effect size observed in GWAS, with small effects for frequently occurring variants and increasingly strong effects for rare variants (0.5%<MAF<1%) had stronger effects. For example, rs11591147 coding for a low-frequent (MAF=1.8%) amino acid substitution in *PCSK9* (p.R46L) was the most strongly associated variant, associated with a 0.13 s.d. value increase per T-allele (p<6.5×10^−65^) with embedding92, one of the polygenic embeddings.

**Figure 2.**
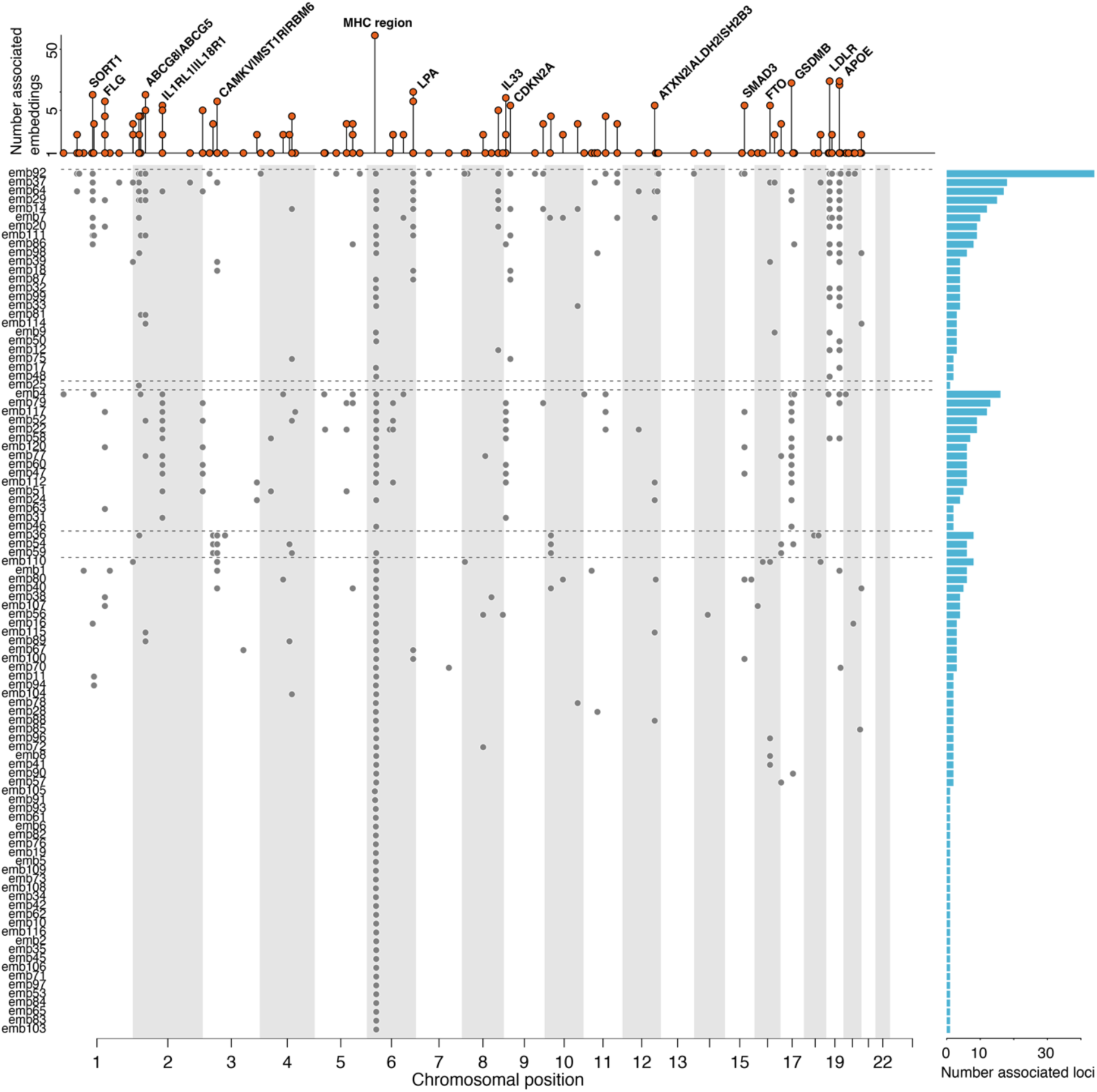
Embedding-associated lead credible set variants. The main panel displays significantly (p<5×10^−8^) associated lead credible set variants according to the genomic position and separated by embedding (‘emb’). For displaying purposes only, embeddings were clustered based on shared associations (r^2^≥0.8) using Louvain clustering with edge weights. The resulting five clusters are indicated by dashed lines. The top panel shows the number of associated embeddings per locus, aggregating proximal variants by LD (r^2^≥0.8) and candidate genes are given for pleiotropic loci. The right panel displays the number of independently associated loci per embedding. Results are based on linear regression models and associated statistics can be found in **Supplementary Tab. 3**.

A total of 100 lead credible variant – embedding associations had a posterior inclusion probability (PIP) >0.5 indicating high confidence in the underlying causal variant. For example, rs13107325 (MAF=7.4%), a missense variant overlapping *SLC39A8* (p.A391T), inversely associated with embedding89 (beta = -0.02; p-value<9.9×10^−11^) was fine-mapped as the candidate causal variant with high confidence (PIP=92.6%). The variant or proxies thereof (r^2^≥0.8) have been reported for >100 traits in the GWAS catalog (**Supplementary Tab. 3**), spanning psychiatric disorders and brain morphology, musculoskeletal disorders, or blood lipids. The critical role of the electroneutral divalent metal cation:bicarbonate symporter encoded by *SLC39A8* for the cellular uptake of essential trace metals such as zinc, manganese, or selenium, likely explains these pleiotropic effects across disease domains and the associations with surrogates of disease trajectories in our study.

We observed considerable sharing of genetic variant associations across embeddings, partially resembling observational correlation analysis. Among 151 independent credible sets, referred to hereafter as loci identified across all embeddings by clumping coinherited variants (r^2^>0.7), more than a third (n=54) associated with two or more embeddings (**Fig. 2**). This included five credible sets associated with ten or more embeddings. All of which tagged regions of established pleiotropy according to the GWAS Catalog^18,19^, but almost exclusively for diseases either related to lipid metabolism, such as multiple variants at the *APOE* locus (e.g., rs429358 associated with 26 embeddings), or immune system associated respiratory diseases and malignancies (e.g., rs796074792 associated with 14 embeddings).

In contrast, strong and specific associations across embeddings at loci included, amongst others, rs1894692 (MAF=2.2%) downstream of *F5* associated with lower values of embedding1 (beta=- 0.03; p-value<5.9×10^−18^). The locus is known to associate with coagulation defects and associated disorders aligning with a putative effect on factor V, a central regulator of haemostasis, encoded by *F5*.

### Sex difference in the embedding space rarely translated into sex-specific genetic findings

Despite the pronounced sex separation of embedding values (**Fig. 1g**), we observed only 20 locus– embedding associations with significant evidence (p<5×10^−8^) of heterogeneity in effect estimates based on sex-stratified genome-wide association testing (**Supplementary Tab. 4**). Most (n=13) of the associations showed evidence for sex-specific effects (n=4 female only; n=9 male only; defined as p<5×10^−8^ in one and p>0.05 in the other sex), including four loci also identified in the sex-combined analysis. For example, rs4007642 (MAF=48.4%) showed male-specific associations with embedding14 (beta_men_=-0.02, p-value_men_<1.4×10^−23^; beta_women_=-0.003, p- value_women_=0.43) and embedding87 (beta_men_=-0.02, p-value_men_<2.2×10^−22^; beta_women_=-0.003, p- value_women_=0.15). The variant or proxies thereof have been repeatedly linked to various diseases, spanning cardiovascular diseases, metabolic diseases, but also cancers (**Supplementary Tab. 3**), and maps to the *CDKN2A/B* gene cluster encoding for cellular senescence genes a hallmark process of age-associated disease onset^20^. We observed only one variant on the X-chromosome with sex-specific effects, with rs138669696 being associated with embedding48 in women only (beta_men_=0.005; p-value_men_=0.05; beta_women_=-0.02; p-value_women_<1.4×10^−8^). Collectively, these results indicated that, like previous disease-specific studies^21^, pronounced sex-differences in embedding values cannot be explained by common germline genetic variation.

### Effector genes implicate cholesterol metabolism and IL-1-family alarmin signalling as shared mechanisms across embeddings

Since most identified variants resided in non-coding regions of the human genome with no clear immediate role in (disease) biology, we used an ensemble classifier integrating various layers of functional genomic information to link variants to effector genes. We successfully assigned a single candidate effector gene for 332 out of 434 lead credible variant – embedding associations with high confidence (score>0.25; **Supplementary Tab. 3**) and subsequently subjected those to pathway, tissue, and cell type enrichment testing.

We observed 15 embeddings with at least one significantly enriched pathway (FDR<5%), that segregated into three groups based on pathways shared across embeddings (**Supplementary Tab. 5**). The first group comprised five embeddings enriched for effector genes in interleukin-1 (IL-1)- family signalling (e.g., embedding60: 73-fold; FDR<7.4×10^−5^) and resembled the epithelial-alarmin axis (**Fig. 3a**). Across embeddings, we recurrently mapped the IL-1-family alarmins and their receptors, *IL33* (ten embeddings), *IL18R1* (six), and *IL1RL1/ST2* (six), together with the epithelial cytokine TSLP (rs1837253, PIP=1.00), the 17q21 *GSDMB/ORMDL3* locus (ten), *SMAD3* (six), and *LRRC32* (four), recovering the canonical type-2 inflammatory loci of asthma and allergic diseases^22^. A second group (seven embeddings) was strongly enriched for effector genes involved in cholesterol metabolism and plasma lipoprotein clearance (e.g., embedding92: 52-fold; FDR<1.1×10^−13^), tagging most established druggable targets but also emerging ones such as *LPA* (see below; **Fig. 3b**). The remaining three embeddings were enriched for distinct pathways, including, a 51-fold enrichment for effector genes involved in ‘signalling by ROBO receptors’ (embedding36; FDR<1.3×10^−4^).

**Figure 3.**
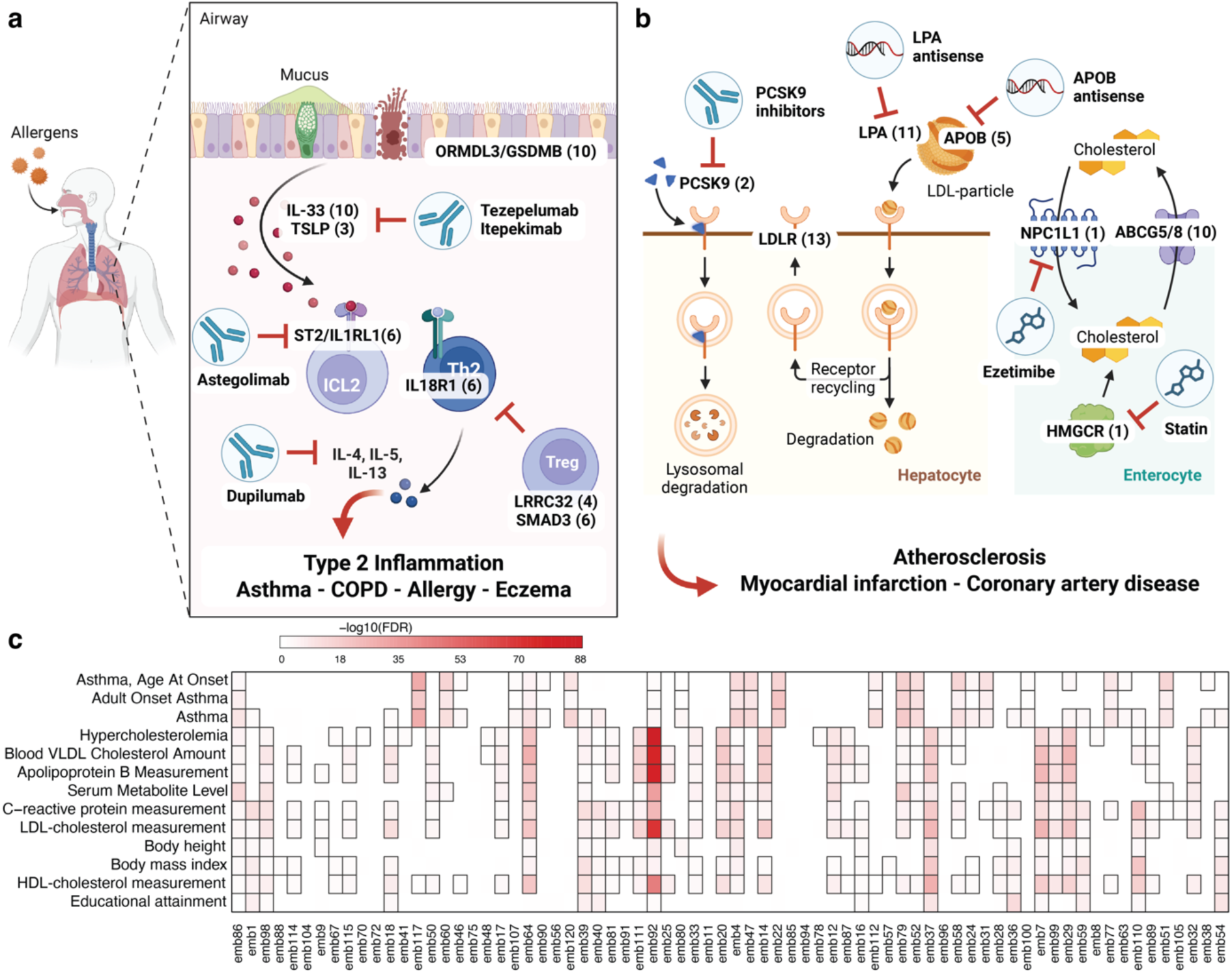
Embedding associated loci are enriched for those linked to specific disease risk. **a** Candidate effector genes recovered across embeddings reconstruct the epithelial-alarmin-type-2 inflammation pathway. Node subtitles give the assigned effector gene and the number of embeddings at which it was associated. Drugs were added based on colocalization evidence. **b** Effector genes recovered across embeddings reconstruct the cholesterol and lipoprotein-clearance axis. Same logic of assignment as for **a**. **c** Results from enrichment testing (Fisher’s test) of embedding associated loci among those previously reported for outcomes curated in the GWAS Catalog (y-axis)^18^. The colour gradient indicates the significance of enrichment following correction for multiple testing using the false discovery rate (FDR). Embedding – outcome pairs reaching significance are highlighted by frames. Findings have been collapsed to reduce redundancy and the full results can be found in Supplementary Table 8. **a** and **b** were created with BioRender.com

These results were closely mirrored by tissue and cell type enrichments (**Supplementary Tab. 6- 7**). For example, effector genes associated with embedding92 were enriched for expression in liver (odds ratio (OR): 17.7; FDR<1.2×10^−9^), duodenum (OR: 11.5; FDR<2.1×10^−4^), and small intestine (OR: 9.1; FDR<1.9×10^−3^), the major sites of lipoprotein metabolism. Similarly, genes with enhanced expression in brain excitatory and inhibitory neurons were 19-fold enriched for effector genes linked to embedding36, cell types that rely on ROBO receptor mediated signalling during development and axon guidance^23^ and hence tagging a domain of brain and mental health.

### Embedding associated loci are enriched for susceptibility to asthma and cardiometabolic diseases

All identified loci or proxies thereof (r^2^>0.1) have previously been reported in the GWAS Catalog to be associated with various traits and diseases^18^, including 131 loci likely tagging the same underlying causal variant (r^2^>0.8; **Supplementary Tab. 3**). Results that supported the biological plausibility of our findings and allowed us to refine the understanding of the domains of health learned by the Delphi-2M model.

We observed significant (false discovery rate (FDR) <5%) enrichment of previously reported disease and complex trait associations among identified loci for 51 of the embeddings (**Supplementary Tab. 8**) indicating that those represented known domains of health. Notably, including traits and risk factors that were never used to train Delphi-2M. Loci shared across embeddings thereby drove most of the enrichment and identified hypercholesterolemia (e.g., 1243-fold enrichment of loci among those associated with embedding92; FDR<1.7×10^−86^; enriched among 32 embeddings) and asthma susceptibility (e.g., 77-fold enrichment among loci associated with embedding4; FDR<8.0×10^−16^; enriched among 33 embeddings in total) as the most strongly and frequently enriched trait categories (**Fig. 3a-c**). Body composition (e.g., 65-fold enrichment of loci for body mass index among loci associated with embedding59; FDR<1.7×10^−9^; n=31 embeddings) and educational attainment (e.g., 47-fold enrichment among loci associated with embedding36; FDR<7.6×10^−13^; n=13 embeddings) further emerged as major domains with shared enrichment profiles across embeddings, reflecting two of the most important modifiable factors of disease risk (**Fig. 3c**). We note, that while we observed enrichment of GWAS traits, most heritable embeddings likely represented a convolution of different domains of health.

We corroborated GWAS Catalog results by observing that 123 out of 144 embedding associated loci outside the MHC region were significantly (p<5×10^−8^) associated with the occurrence of at least one ICD-10 coded diagnoses in UKB^24^, that is the data used to train Delphi-2M (**Supplementary Tab. 9**). This included 13 loci with strong (|odds ratio| > 1.5) effects on the risk of diseases such as rs75331444 (MAF=6.52%), intronic of *ABCG8,* associated with an almost 2- fold increased risk for cholelithiasis (odds ratio=1.88; p-value<10^−300^).

However, we also observed five loci not associated (p>0.05) with any ICD10-coded disease in UKB illustrating genetic findings learned by the model that could not have been discovered using the input data. All but one (tagged by rs548921462) have already been reported in the GWAS Catalog to be associated with, amongst others, blood lipid levels (chr11:116623213:TA>T associated with three embeddings), body mass index and substance abuse (chr16:28501781:TA>T associated with embedding110), but also UKB participation bias (e.g., chr3:23638844:CA>C associated with embedding92).

### Embedding loci recover approved drug targets and map druggable axes of lifetime morbidity

Genetic variation that mimics a drug’s mode of action can provide an in-human readout of the effect of pharmacological modulation, and hence we next asked whether embedding loci pointed to actionable biology. To this end, we systematically tested a curated set of 426 genetic loci proxying the effect of 724 approved or clinically advanced (phase 3+) drugs, acting through the drug’s canonical target, as a ligand - receptor pair, or as part of a protein complex for evidence of colocalisation with embedding associated loci (posterior probability of a shared causal variant ≥80%; see **Methods**).

Across 98 embeddings, we observed evidence for drug-mimicking effects at 71 loci, implicating 55 candidate causal genes proxying 81 canonical drug targets, for a total of 217 distinct drugs across 36 indications (411 embedding - locus associations in total; **Supplementary Tab. 10**). Notably, while recovering more drugs in general (n=463), twelve drugs were not recovered when implementing the same pipeline across GWAS summary statistics for 762 ICD-10 coded diagnoses that served as input for Delphi-2M. For example, we observed strong evidence for colocalization (PP=98.3%) at a locus proxied by rs11811788 between embedding84 and atopic dermatitis. The variant likely acts via *TNFSF4* encoding for the cytokine tumor necrosis factor superfamily member 4 (OX40) the receptor being the target of Rocatinlimab that recently proved effective against atopic dermatitis^25^.

Druggable findings segregated into partially distinct therapeutic areas along associated embeddings (**Fig. 4a**) comprising cardiovascular, metabolic/endocrine, immune, neurologic, and oncologic indications. About a fifth (n=19) of embeddings associated with 5 or more such drug-mimicking loci (range: 1-12; **Fig. 4a**). Conversely, 21 drugs mapped to ≥18 embeddings (range: 1-35; examples **Fig. 4b-c**) partly enabled through protein complex partners, like 33 colocalising associations at the *LDLR* locus implying Tafolecimab, a proprotein convertase subtilisin/kexin type 9 (PCSK9) inhibitor that blocks degradation of the low-density lipoprotein receptor encoded by *LDLR* (**Fig. 3b**).

**Figure 4.**
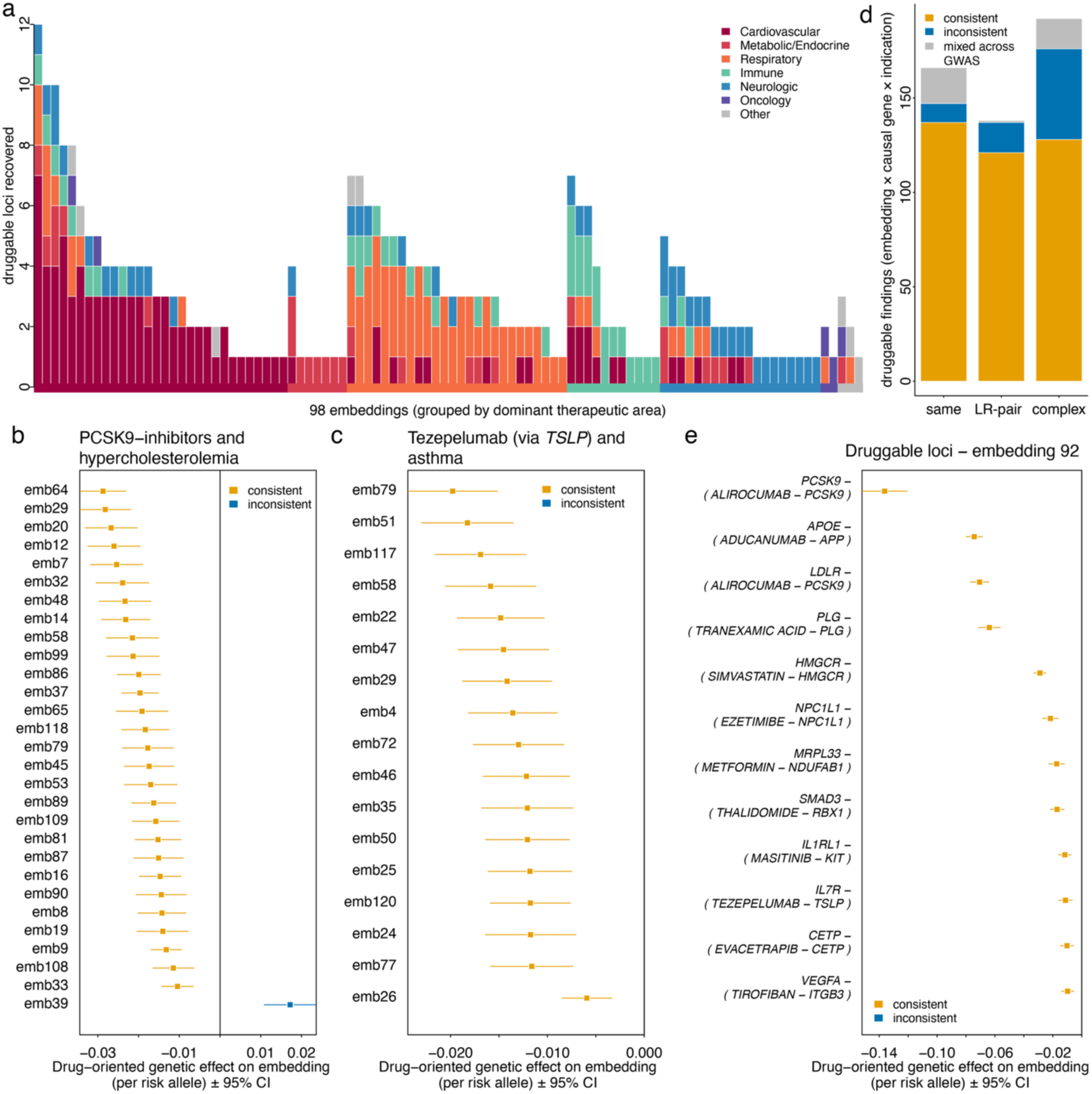
Genetically inferred druggable domains of lifetime morbidity proxied by patient embeddings. **a** Summary of loci associated with one of 98 embeddings that mimicked the action of a drug. That is, the embedding associated variant was also associated with the indication of an approved or advanced (phase III) drug (posterior probability of same variant ≥80%) and acted either through the canonical target, as ligand-receptor (LR) pair, or as part of a protein complex. Colouring indicates therapeutic areas for recovered drug effects and embeddings were ordered by the number of ‘druggable loci’ and composition of therapeutic areas. **b-c** Forest plot (linear regression estimates) for example loci illustrating shared effects across embeddings for the two most promiscuous loci mimicking PCSK9-inhibitors (**b**; via rs142130958) and Tezepelumab (**c**; via rs1837253). **d** Summary of effect directions at druggable loci. Since embeddings have no natural direction, we oriented effects based on matching phenotypic associations for the colocalising drug indication. Yellow indicates effects consistent with beneficial effects on indication and embedding, whereas blue indicates opposing, potentially harmful effects. **e** Genetic effects (linear regression analysis) of variants mimicking drug action across 12 loci on levels of embedding 92. Loci and associated drugs with target genes (in brackets) are annotated. Some may act indirectly.

Because embeddings carry no natural orientation, such as disease increasing or decreasing, we oriented each genetic effect against the phenotypic association of the colocalising indication. That is, whether participants with a recorded indication had higher or lower values of the colocalizing embedding. In doing so, we classified every embedding-target-indication finding (n=496) as directionally consistent with a beneficial drug effect, potentially adverse, or directionally inconsistent across independent GWAS for the same indication (**Fig. 4d**). Most findings were consistent (n=386; 78%), with 75 (15%) potentially adverse and 35 (7%) mixed. Among unambiguous findings, consistency increased with mechanistic stringency, from 72% at protein-complex loci to 88% for ligand-receptor pairs and 93% at canonical targets. Inconsistent and maybe potentially adverse findings were concentrated at complex-partner loci (n=49; 65%), where the colocalising causal gene was not the drug’s canonical target, e.g., anti-amyloid antibodies tagged by the *APOE* rather than the *APP* locus. We note that, because orientation was anchored to each drug’s own indication, a consistent direction likely recapitulated the established effect of the drug on that indication and an independent effect on the broader healthcare trajectory remains to be proven.

Finally, our drug matching exercise also corroborated the observation that embeddings cover diverse aspects of health. For example, embedding 92 mapped to 12 drug-mimicking loci (**Fig. 4e**) capturing all currently approved lipid-lowering therapies (statins (*HMGCR*), ezetimibe (*NPC1L1*), CETP inhibitors and *APOB*- and PCSK9-targeting agents) along with medications approved for severe asthma control, platelet activation inhibitors, and other immunomodulatory drugs. Conversely, drug-mimicking loci shared across embeddings captured immunomodulatory therapy (**Fig. 3a**). The alarmin-axis loci colocalised with targets of severe-asthma and chronic obstructive pulmonary disease (COPD) biologics such as Tezepelumab (anti-TSLP; 22 embeddings), Itepekimab (anti-IL-33; 18), and Astegolimab (anti-ST2/IL1RL1; 18).

### Comorbidity and a heritable cardiometabolic state independently explain predictive accuracy

Having shown that Delphi-2M’s embeddings implicitly encode heritable risk factors it was never supplied as inputs, we next quantified what kind of disease knowledge underlies its predictive accuracy, that is, direct disease – disease relationships or those mediated through shared risk factors (**Fig. 1a**). To separate these two axes, we associated each disease’s area under the receiver-operating curve (AUC), as achieved by the model on held-out participants, to two complementary descriptions of its context: 1) its node characteristics in a partial-correlation network across 780 disease tokens (n≥100 cases), and 2) the single-exposure discrimination gain (ΔC-index over age and sex) of 148 measured risk factors (spanning lifestyle, diet, body composition, inflammation, frailty, circulating biomarkers, and blood cell counts) in Cox-survival models, capturing putative common causes the model may have learned implicitly (**Fig. 1b**). In other words, we tested whether per disease AUC values associated with disease characteristics derived from a co-occurrence network and systematic association testing totalling 276 features.

We identified four independent features that associated with and explained variation in per disease AUC values, two mapping to each axis, using Bayesian-fine mapping for feature selection (**Fig. 5a-d**). The number of significantly co-occurring diseases (beta=0.65; p-value<6.7×10^−46^) and to a lesser extent their median partial correlation coefficient (beta=-0.11; p-value<2.9×10^−2^) supported the comorbidity axis, whereas the predictive value (ΔC-index) of information on long-standing illnesses (beta=0.34; p-value<1.5×10^−17^) and falls in the last year (beta=0.14; p-value<7.1×10^−4^) proxied a complementary, likely cardiometabolic/inflammatory axis. Annotation of the latter axis was supported by discrimination gains of total cholesterol (beta=0.27, p-value<3.0×10^−14^), C-reactive protein (beta=0.27, p-value<7.0×10^−13^), and waist-to-hip ratio (beta=0.26, p-value<2.2×10^−12^) themselves being associated with accuracy and tightly correlated to the frailty features (r≥0.5), mirroring the lipid and inflammatory genetic enrichment of embeddings (**Fig. 5e-g; Supplementary Tab. 11**).

**Figure 5.**
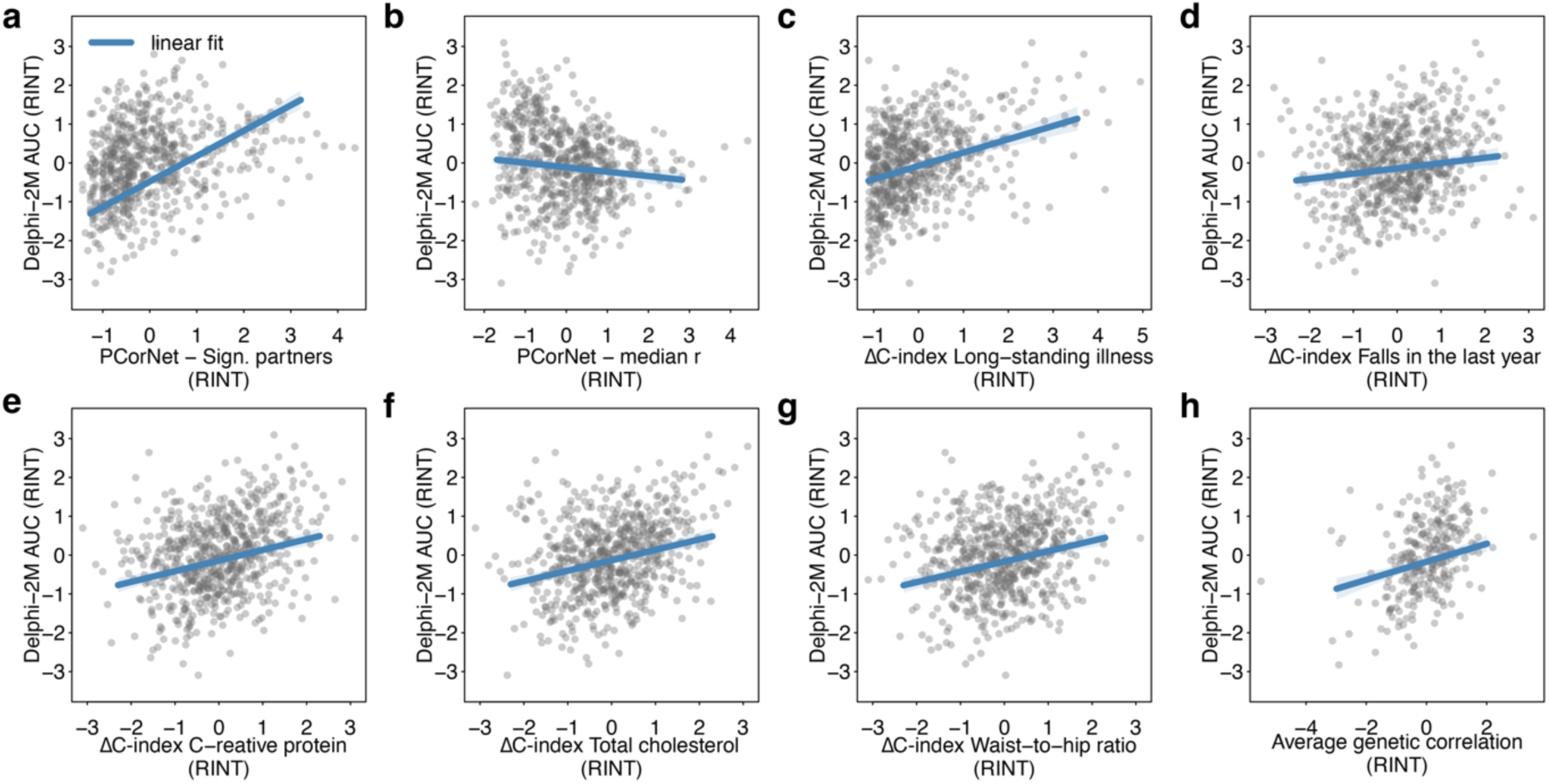
Predictive capacity of Delphi-2M extends beyond a learned comorbidity structure. **a-g** Scatter plots opposing area under the curve achieved by Delphi-2M across ICD-10 codes averages across age- and sex-stratified bins on a ten-year horizon against: **a-b** Measures derived from a partial correlation network (PCorNet) across lifetime disease co-occurrence based on the entire UK Biobank cohort, including the number of significant partners (sign. partners; **a**), i.e., the higher the comorbidity burden of a disease the better Delphi- 2M can predict it, and the median partial correlation coefficient (**b**); **c-g** Changes in C-index over an age, sex, and centre baseline from Cox-proportional hazard models including each of the named variables on the x-axis as single exposure. **h** Similar scatterplot but opposing to the average significant genetic correlation of a given ICD-10 coded diagnoses with all other diagnoses. All measures were rank-based inverse-normal transformed (RINT) to ensure comparable scales across features. Linear fits are based on linear regression models adjusting for the case counts for each diagnosis (c-g) and additionally for factors displayed in **a-d** for **h**.

Prediction accuracy was also associated with the average significant genetic correlation of a disease with all other diseases (**Fig. 5h**). An association that persisted after adjustment for both the comorbidity and cardiometabolic axes in multivariable linear regression models (beta=0.21; p-value<8.1×10^−7^) (**Fig. 5h**), indicating that shared genetic liability carries information on predictability not captured by the measured comorbidity structure.

### Delphi-2M has learned the predictive cross-disease value of most routine biomarkers and plasma proteins, and improvement is restricted to disease-specific markers

Having shown that the embeddings encode both a comorbidity and a heritable cardiometabolic/inflammatory domain, we finally asked which (blood) biomarkers may improve risk prediction or are already implicitly learned by the model. Across 117 routine biomarkers and continuous measures of risk, the median within-sex variance explained (R^2^) by the 120 embeddings was 0.06 (IQR 0.03 - 0.15), and only 21.3% reached R^2^≥0.2. We observed even lower results across 2,919 plasma proteins with a median R^2^ of 0.02 (maximum 0.43; 2% ≥0.2; **Supplementary Tab. 12**) (**Fig. 6a**). Well explained risk factors by embeddings were restricted to measures of adiposity and body composition (R^2^: females - median 0.37, IQR 0.24 - 0.46; males - median R^2^ 0.32, IQR: 0.23 - 0.38), and proteins that represent process of ageing, such as EDA2R^26,27^ (R^2^: women=0.39, men=0.44), and general illness, such as GDF-15^28^ (R^2^: women=0.39, men=0.46), rather than any single organ marker. Those results implied that the model likely learned the broad, densely recorded dimensions of metabolic and ageing-related risk, but not the specific biochemical state captured by most individual markers, such as specific markers of organ health or alike.

**Figure 6.**
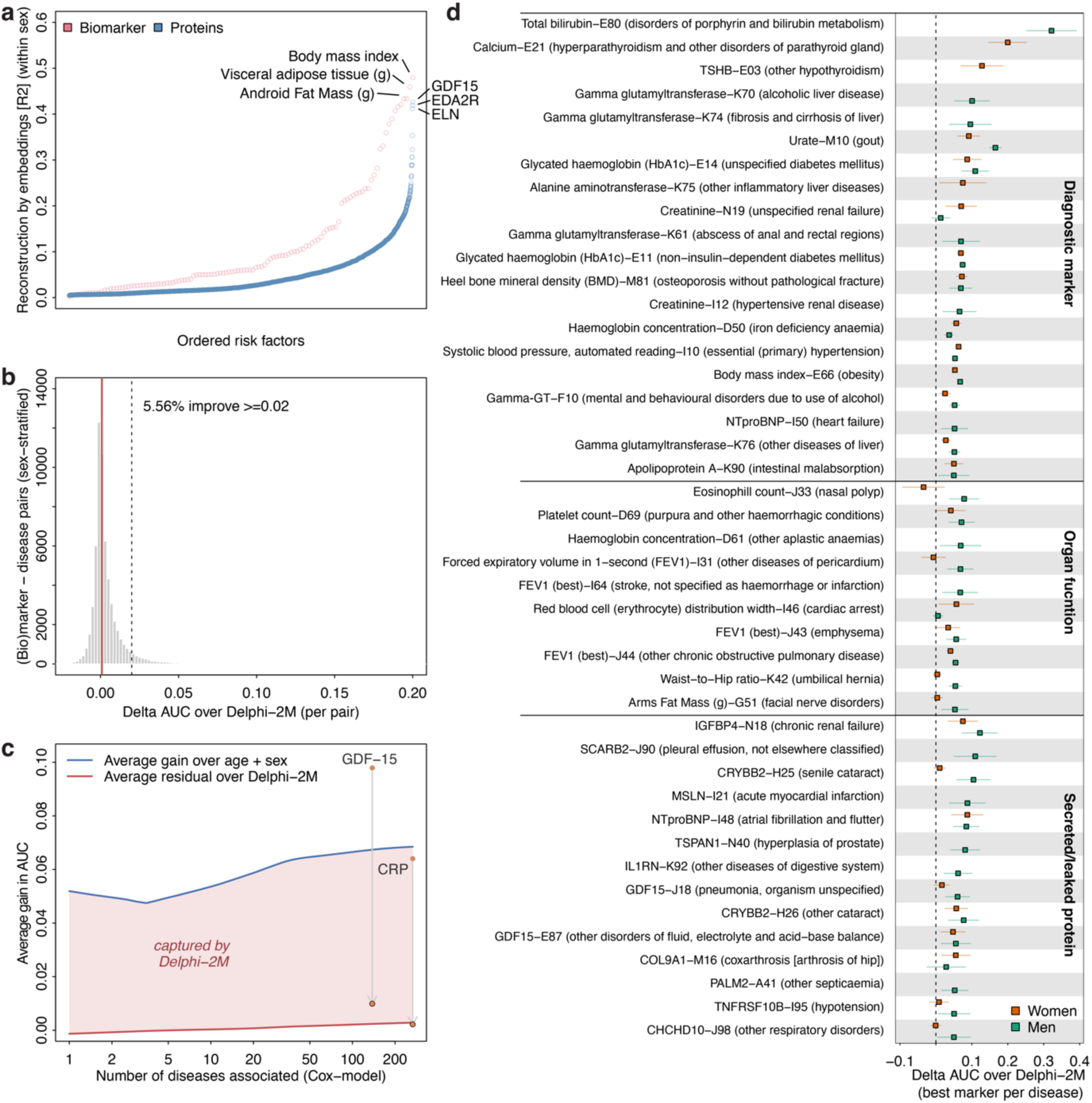
Internalisation of and improvement by biomarkers of the Delphi-2M model. **a** Explained variance achieved by regressing all 120 embeddings on (protein) biomarker levels using linear regression models. Results were averaged across sexes from within sex predictions. **b** Histogram for the improvement in area under the receiver operating curve (AUC) over and above a linear predictor derived from Delphi-2M across 59,749 biomarker – disease pairs (30,164 in women, 29,585 in men) that showed significant (p<3.3×10^−6^) associations in Cox-proportional hazard models. AUCs were computed based on a 10-year horizon in cross-validated out-of- sample predictions in age-stratified sets. **c** Association between biomarker pleiotropy (number of significantly associated diseases in Cox-models) and achieved AUC gain, with (red) and without (blue) adding Delphi-2M’s linear predictor for the respective disease. The red area indicates the predictive gain likely captured by Delphi-2M. **d** Selected biomarker – disease examples that improved the AUC over Delphi-2M ≥ 0.05. Examples were manually grouped based on possible mechanisms. Improvement in AUC is shown by sex and confidence intervals are based on cross-validation. Combinations missing either sex were due to low case numbers in the validation set (n≤100).

Despite the generally low explained variance, adding a measured marker to Delphi-2M did not improve prediction for most outcomes (**Fig. 6b**). Over 21,574 biomarker - disease and 38,175 protein - disease pairs with at least 100 incident cases and significant (p<3.3×10^−6^) associations in survival models, the median gain for biomarkers and proteins over age and sex was substantial (median of 0.044 and 0.063, respectively, in 10-year AUC-improvement estimated from out-of-sample by cross-fitting within age and sex strata), yet the median gain over the same baseline plus Delphi-2M was 0.001 for both (**Fig. 5b; Supplementary Tab. 13**). That is, roughly 98% and 99% of the predictive value of the tested risk factors and proteins above age and sex was already captured by the disease trajectories learned, and only about 4% of biomarker pairs and 7% of protein pairs improved the AUC ≥0.02 (**Fig. 6b**).

This redundancy was not uniformly distributed across tested markers but scaled with how broadly a marker associated with disease onset. Markers associated with more diseases were stronger predictors over age and sex (Spearman ρ between pleiotropy and gain over demographics = 0.89 for biomarkers and 0.41 for proteins), but because their median residual gain over Delphi-2M stayed close to zero, the model already learned almost entirely that additional value without being ever supplied this biomarker (**Fig. 6c; Supplementary Tab. 14**). A prominent example was GDF-15, the most pleiotropic protein measured (associated with 141 and 135 diseases in women and men, respectively) and among the strongest predictors over demographics (median ΔAUC = 0.098), yet 90% of its predictive value was captured by Delphi-2M (median residual ΔAUC = 0.010) likely explained by the embeddings ability to reconstruct its risk-associated fraction. The next most pleiotropic proteins behaved similarly (e.g., TNFRSF10B, PLAUR, ADM, and IGFBP4: 94 – 113 associated diseases and 89 - 96% gain already captured). Since a longitudinal health record reconstructs accumulated morbidity well, the proteins that integrate the most morbidity are those a trajectory model best approximates, consistent with Delphi-2M having learned shared, multi-disease risk rather than organ-specific biochemical states.

We lastly observed substantive improvements (ΔAUC≥0.05) for 44 out of 301 commonly occurring diseases (n≥100 cases) tested separately across the sexes (**Fig. 6d; Supplementary Tab. 15**). These segregated into three different classes: 1) established diagnostic biomarkers, such as total bilirubin for the onset of bilirubin disorders (ICD-10 code E80 in men; +0.32 AUC; explained variance by embeddings R^2^=0.03), 2) direct measures of organ function, such as forced expiratory volume in 1-second (FEV1) for the onset of chronic obstructive pulmonary disease (ICD-10 code J44; +0.05 in men; R^2^=0.28), and 3) tissue-specific secreted/leaked protein biomarkers, such as NT-proBNP and the onset of atrial fibrillation (ICD-10 code I48, +0.08 AUC for each sex; R^2^: females=0.14, males=0.28). Collectively, these results further corroborated findings from genetic analyses in showing that Delphi-2M has implicitly learned shared risk factors predisposing to multiple diseases and outlines areas of putative improvement when supplementing the model with disease-specific biomarkers.

## DISCUSSION

Foundation models that forecast disease onset based on prior medical records of a patient are gaining traction with the promise to advance personalised medicine^1,6^, but which concepts they have learned to predict onset can often not be inferred sufficiently well from input data^29^, raising concerns about their adoption in healthcare^30^. Here, we show that the latent representations (embeddings) of an EHR-derived foundation model comprised a reproducible, heritable structure, and that genetics can identify implicitly learned concepts not inferable from input data. By performing genome-wide association testing on Delphi-2M’s embeddings, we 1) demonstrated significant SNP-based heritability for all embeddings and identified 434 embedding – locus associations across 151 loci and 98 embeddings, 2) assigned candidate effector genes that converged on cholesterol metabolism and the IL-1-family epithelial-alarmin pathway, 3) showed that embedding-associated loci were strongly enriched for blood lipids, body composition, and asthma and recovered, amongst others, the targets of essentially all approved lipid-lowering and severe-asthma therapies, and 4) established that the embeddings captured the cross-disease predictive value of many routine biomarkers and plasma proteins, with improvement in single biomarker settings restricted to disease-specific markers. However, our findings also pointed to potentially learned biases, including participation in UK Biobank and implicitly learned concepts of socioeconomic status and educational attainment. We believe that these findings demonstrate the unique ability of germline genetic studies to surface implicitly learned concepts of otherwise ‘black box’ foundation models trained on EHRs.

Feature attribution techniques try to reconcile, how input tokens are used by the model to explain prediction^7,31^ but largely fall short in quantifying the implicit context the model has inferred^32^. Speech is a natural analogy, by which transformer models have implicitly learned the rules of syntax and grammar implicitly from reproducing text. While such learned rules can be easily verified in newly generated text, this represents a fundamental challenge to any clinical application, such as risk forecasting or treatment decisions. Those require an in-depth understanding of how and why the model reached decisions, the exact context it has learned and based its decisions on to ensure fair and equitable applications. We showed here that genetic association testing of patient-level embeddings, that underpin model predictions, paired with state-of-the-art post hoc analyses can identify implicitly learned concepts that represent biological domains of risk the model was never supplied with. The genetic results thereby supported and augmented current concepts of lifetime multimorbidity: 1) an epithelial-alarmin pathway, the canonical type-2 inflammatory loci of asthma and allergic disease^22^, 2) a blood lipid domain combining LDL-cholesterol clearance but also lipoprotein(a), 3) the cellular-senescence locus *CDKN2A*, and 4) established pleiotropy hubs such as *ABO*, the 12q24 *SH2B3-ATXN2* locus and *GCKR*. We hence propose genetic decoding as a general, model-agnostic complement to feature attribution for understanding which domains of risk health-trajectory models have learned.

Our genetic findings are supported and considerably extended a recent Bayesian model of longitudinal records that, integrating polygenic scores directly, that reported a heritable cardiovascular signature enriched for variants near *LPA*, *APOE*, and *PCSK9*^12^. The independent recovery of the same lipid architecture from an unsupervised foundation model that was never supplied genotypes strengthens thereby the biological plausibility of both. The work by Urbut and colleagues^12^ built an interpretable, genetics-informed model for prediction and discovery, whereas we treated genotype as a strictly external, training-independent domain to explain a model we did not design and whose embeddings carried no genetic input. It is a hence a design choice motivated by the prediction goals, on how genetic data is implemented as data modality, but given that most predictive information is contained in modalities that capture risk dynamically over lifetime, we argue here in favour of the explanatory role.

A substantial number of embedding associated loci coincided with those mirroring the effect of approved and clinically advanced drugs, such as LDL-cholesterol lowering across various targets but also the epithelial-alarmin pathway that underlies severe asthma and now defines its biologic therapies. The recovery of approved drug targets across these pathways is consistent with the enrichment of genetic support among successful therapies^14,33^, but need to be interpreted with caution. Ideally, those findings highlight interventions that change the lifetime disease trajectory of people, such as non-canonical benefits from anti-hypertensive or lipid lowering medication, but our results are currently bound to directional consistency largely recapitulating the established effect of the drug on that indication rather than demonstrating an independent effect on the broader trajectory. The latter also directly hints at potential room for improvement when training and evaluating EHR-based foundation models, with >200 drugs likely to be recovered based on standard single input analysis.

We showed that the embeddings did only explain variance in few selected biomarkers while still capturing the risk associated fraction for most. While some of these findings can be explained by design, i.e., Delphi-2M used body mass index, smoking, and alcohol consumption as input tokens, it has direct implications for biomarker studies. For example, GDF-15 was the most pleiotropic protein in our analysis and among the strongest predictors over age and sex and has been advanced as a near-universal prognostic marker across cardiometabolic disease, cancer, and ageing^14,34^, yet roughly 90% of its incremental predictive value was already learned by the trajectory model. These findings supported the notion of GDF-15 as a non-specific signal of accumulated somatic distress^28^. In contrast, substantive improvement among the common diseases tested were restricted to diagnostic or organ-leakage markers tied to a specific disease, such as NT-proBNP for atrial fibrillation, refining our earlier observation that sparse protein signatures help most through disease-specific predictors^15,35^.

Our results may inform a design principle for the next generation of EHR-based foundation models. Germline genotype has three properties that can aid in how EHRs are combined for prediction: 1) it is fixed at conception, that is, predates any record, 2) it lies outside the process the model observes, that is, the vast majority of health records is allocated not based on genetic information, and 3) causal variants, albeit present at different frequencies, act similarly across populations^36^. Because it therefore cannot be a product of how or where care was recorded, the heritable fraction of a model’s representation, the part that recovered established causal biology, is by construction the fraction least dependent on documentation. We observed both components in Delphi-2M. Predictive accuracy associated with a heritable cardiometabolic/immunomodulatory domain of risk and a disease’s genetic correlation with the wider spectrum, independent of measured comorbidity, whereas other embedding structure reflected ascertainment, loci tagging participation bias rather than disease, and sex differences unexplained by genetic variation. We therefore propose genetic rediscovery, the heritability of learned embeddings and their recovery of known causal loci and risk factors, as an interpretable quality metric and a candidate training or model-selection objective that draws on tens of thousands of studies providing direct interpretational support^18^. Representations optimised for genetic rediscovery must thereby concentrate on population-invariant causal biology, such as protein altering variants, to reduce the documentation-driven biases that threaten equitable performance. We note that this potential is conditional, since genetic reference data remain heavily skewed towards European-ancestry participants^37^, and polygenic instruments transfer poorly across ancestries^38^, so genetic anchoring will improve fairness only if built on multi-ancestry genetics and confident mapping of truly causal variants to not risk entrenching the disparities it could help resolve.

Since the original model weights for Delphi-2M were not available, all embeddings were derived from a model we retrained rather than from the published checkpoint. While our replication aligned closely with performance metrics of the original work it was not identical. However, residual differences were most likely attributable to the underlying UK Biobank data release and to the software and hardware environment rather than to the modelling procedure itself. Importantly, our central conclusions did not rest on exact reproduction of the checkpoint. Because germline genotype was never supplied to the model and is fixed at conception, the heritable fraction of the embeddings is anchored to training-independent biology, and its convergence on established causal loci provides external validation that is, by construction, insensitive to the residual performance gap. We would therefore expect the qualitative genetic architecture we report to be robust to implementation, while acknowledging that the precise embedding values, and hence individual effect estimates, may differ from those obtainable from the original weights.

Several limitations must be considered when interpreting our results and conclusions. First, the embeddings carried no natural orientation, which constrained directional and mechanistic interpretation and required external anchoring of effect directions using linear association testing. Relatedly, the biomarker redundancy we report is measured over a frozen Delphi-2M predictor in a single-marker, single-disease incremental design, and estimated the value of supplementation in that setting rather than the ceiling of these models. The range of diseases we were able to test, that is, common complex rather than rare diseases, also restricted further generalisation of our findings and we have already shown that rare diseases may benefit most from targeted biomarker studies^15^ and supplementation of biomarkers for participants with few EHRs is an open topic. Second, UK Biobank is a volunteer cohort of predominantly European ancestry and is not representative of the general population^39^ and a minority of loci plausibly tagged participation bias rather than biology. Finally, much of the genetic signal recapitulated known disease GWAS signals, so the orthogonality we claim lies in the explanatory potential, a causally anchored, training-independent probe, more than in the discovery of unreported loci. Within these bounds, our findings showed that human genetics can be used to audit and explain what otherwise cryptic EHR-based foundation models have learned, an approach we expect to generalise to other model architectures and including different data modalities.

## ACKNOWLEDGEMENTS

The authors acknowledge the Scientific Computing of the IT Division at the Charité - Universitätsmedizin Berlin for providing computational resources that have contributed to the research results reported in this paper (https://www.charite.de/en/research/research_support_services/research_infrastructure/science_it/#c30646061).

## AUTHOR CONTRIBUTIONS

Conceptualization: WZ, MP

Data curation/Software: WZ, LK, CB, MZ, MK, MP

Formal Analysis: WZ, MP

Methodology: WZ, BW, JCZ

Visualization: WZ, MP

Funding acquisition: CL, MP

Project administration: MP

Supervision: CL, MP

Writing – original draft: WZ, MP

Writing – review & editing: all

## DATA AVAILABILITY

Individual-level data from UK Biobank are publicly available to bona fide researchers upon application (https://www.ukbiobank.ac.uk/).

## CODE AVAILABILITY

The analysis code supporting the findings of this study will be made available on GitHub upon publication.

## METHODS

### Dataset

This study was based on the individual-level electronic health records provided by the UKB (‘first occurrence’). The UKB is a prospective cohort study in the UK consisting of more than 500,000 individuals. All analyses were conducted under UKB application 44448.

### Replication of Delphi-2M

As the Delphi-2M checkpoint trained by the original developers was not readily accessible, we re-implemented Delphi-2M among 501,939 participants with available EHRs and who not revoked consent based on the same first disease occurrence dataset as the original publication. Individual- level EHRs were represented as sequences of time-stamped tokens, containing clinic event, demographic attribute, and lifestyle factors, along with the age at which the event occurred. The training sequences comprised 1,261 unique tokens spanning diseases, sex and lifestyle; relative to the original setting, the ICD-10 codes F83, M09, O01, P52, P71, P78 and CXX were absent. The complete replication procedure including dataset preprocessing and model training, strictly following the pipeline released in the original Github repository^40^, with training parameters identical with those reported in the original study. All implementations were carried out in Python, primarily using the PyTorch framework^41^, and experiments were performed on a single NVIDIA GPU. Although we closely adhered to the original configuration, minor performance discrepancies remained between the original and re-implemented models, most likely attributable to differences in the underlying UKB data release (first occurrence download: 07/05/2021) and in software and hardware environments.

### Performance evaluation of the replicated Delphi-2M

Following the developers’ evaluation pipeline, the replicated Delphi-2M model was assessed with a focus on disease prediction performance, on a subset of diseases and across the full set of diseases quantified using the area under the receiver operating curve (AUC). In addition, a series of performance visualisations was generated, including analyses of the relationship between the number of tokens in the training set and the corresponding AUC, and AUC distributions stratified by disease chapter and by sex. The effectiveness of the replication was further examined through systematic comparison with the performance reported for original models. Specifically, performance consistency between the two models was quantified by computing the Pearson correlation coefficient on AUCs across all diseases, as well as on the AUC scores aggregated by diseases chapters and by sex. Furthermore, the latent token representations learned by the replicated model were investigated by projecting them into two-dimensional space using UMAP, and the intra- and inter-diseases cluster distances were compared visually between the replicated and original models.

### Individual-wise latent representation generation

The replicated Delphi-2M model was used as an encoder to compute hidden representations for these individuals without further training. Individual health trajectories were represented as sequences of tokens and corresponding ages using the same strategy as during training. The final layer of the model outputs a sequence of hidden states that collectively capture the model’s internal representation of an individual’s health trajectory across all time steps. Individual-level embeddings were generated by mean-pooling the final-layer hidden states across all non-padding tokens, yielding a 120-dimensional embedding vector describing the entire health trajectory of each participant.

### GWAS analysis on individual-wise latent representations

Individual-level embedding generation provided a dataset of 120 features and we restricted the dataset to 441,189 participants of genetically inferred British-European ancestry for genetic analyses. Each embedding was inverse-rank normal transformed before genome-wide association testing (GWAS). The Haplotype Reference Consortium-imputed genetic data, covering all autosomal chromosomes and the X chromosome were used. GWAS were implemented under the additive model using REGENIE (v4.0) through a two-step procedure to account for population structure. High-quality genotyped variants for step 1 were selected by applying the following quality control criteria: MAF>1%, MAC>100, missingness rate<10%, *P_H_*_W*E*_ > 1 × 10^−15^. Variants were further pruned for LD using a 1,000-kb sliding windows, shifting by 100 variants and excluding those with pairwise *r*^2^ > 0.8. Genome-wide predictions were generated from these filtered variants in step 1 using the leave-one-chromosome-out approach and used in step 2 for variant-level association testing. All models were adjusted for age, sex, and the first ten genetic principal components. Association testing was restricted to variants with MAF>0.5%, yielding approximately 14.1 million variants in British-European individuals.

Furthermore, to ensure the reliability and comparability of our results, we harmonised our GWAS results against UK Biobank whole-genome sequencing (UKB WGS) GWAS summary statistics across 763 studies^24^ based on shared rsIDs, retaining variants present in both datasets for downstream analyses. This resulted in 11,893,250 of 14,081,025 tested variants being retained (84.5%).

### SNP-based heritability estimation

We used LD-score regression to estimate SNP-based heritability for the GWAS of all 120 embedding dimensions^42^. Summary statistics for each embedding were reformatted to LDSC-compatible format using munge_sumstats.py^42^, restricting to HapMap3 SNPs via the w_hm3.snplist reference. Heritability was then estimated using ldsc.py.

### Regional clumping and fine-mapping

For each embedding phenotype, we performed regional clumping by generating ±500 kb windows around the variants that exceeded the predefined genome-wide significance threshold, with overlapping windows subsequently merged to define distinct genomic regions using BEDtools (v2.30.0)^43^. The extended MHC region on chromosome 6 (25.5–34.0Mb) was treated as a single region. Then for each genomic region, we selected the genome-wide significant (p<5×10^− 8^) variant with the largest absolute Wald statistic, defined as |*β* ∕ *SE*|, as the regional sentinel variant. This resulted in one sentinel variant per significant genomic region for each embedding.

For each region of interest outside the MHC region, we applied statistical fine-mapping across all associated embeddings using the Sum of Single Effects (SuSiE) model implemented in the susieR R package (v0.14.2)^44^. Briefly, SuSiE applies a Bayesian variable-selection framework within a multiple-regression setting to decompose association signals into credible sets, each expected to capture a single causal variant. All default priors and parameter settings were retained, except that the minimum absolute correlation threshold was set to 0.1. For computational efficacy, we ran SuSiE based on GWAS summary statistics together with an in-sample LD reference panel via the susie_rss() function, following the authors’ recommendations^44^. To decide the number of credible sets within each region, we iterated over the maximum number of single effects parameter in SuSiE from 2 to 10, generating fine-mapping results constrained to a range of maximum numbers of independent association signals. For each run, credible sets with correlated lead variants from distinct sets (*r*^2^ ≥ 0.25) were excluded, and the model with the largest number of credible sets was selected.

We subsequently extracted lead credible sets variants as those with the highest posterior inclusion probability (PIP) within each independent credible set. For loci with multiple credible sets, we jointly modelled the association of all lead credible set variants using a multivariable linear regression model to test for independence, adjusting for age, sex, and the first ten genetic principal components. Credible sets were kept only when the lead variant remained genome-wide significant in both marginal and joint analyses, showed concordant effect direction, and had a joint effect estimate within 25% of the marginal estimate. Credible sets failing these criteria were excluded from the final fine-mapped signals, while variant-level PIPs were preserved for completeness.

### Causal gene assignment

For each lead variant and proxy variants in strong linkage disequilibrium (r^2^>0.5), we queried multiple resources to support effector gene assignment. These included data from the ABC Atlas, fine-mapped eQTL catalogues, Hi-C–based chromatin interaction studies, OMIM^45^, Open Targets^46^, and all annotations used in the FLAMES effector gene annotation framework^47^. In addition, we incorporated predicted functional consequences derived from the Ensembl Variant Effect Predictor (VEP) and CADD scores^48^. We trained an XGBoost model using three complementary variant-to-effector-gene datasets: (i) trans-pQTLs linked to ligand–receptor or protein complex pairs^49^, (ii) the Open Targets gold-standard variant-to-gene dataset^50^, (iii) metabolite QTLs^51^, (iv) ExWAS implicated non-coding credible sets used in FLAMES or PoPs^52^, collectively capturing diverse biological modalities. Finally, effector gene scores were assigned by summing predictions across all five classifiers. Where aggregate evidence was sufficient (maximum classifier probability >0.25), the gene with the lowest combined rank across median and maximum probability was reported as the putative effector gene. Where no classifier exceeded this threshold, the nearest protein-coding gene by genomic position was assigned as a fallback.

### Functional enrichment analysis

We performed pathway enrichment analysis on the effector genes of each embedding with at least one assigned effector gene. For variants lacking an HGNC symbol, the nearest gene was used, while for variants mapped to multiple candidate genes, the gene with the highest median probability was taken as representative. The unique set of effector genes per embedding was queried against the REACTOME, KEGG and CORUM databases through g:Profiler^53^, with the organism set to Homo sapiens. Multiple testing was controlled using the false discovery rate (FDR).

### Heterogeneity analysis to identify sex-differential SNPs

We performed sex-stratified GWAS and assessed between-sex heterogeneity through fixed-effects meta-analysis using METAL (v.2020-05-05)^54^, to investigate the contribution of common genetic variation to the observed sex differences across the 120 Delphi-2M-derived embeddings. GWAS were conducted separately in males and females using REGENIE as described above across chromosomes 1–22 and X, and chromosome-wise summary statistics were concatenated into a single genome-wide file per embedding per sex. For each embedding, the female- and male-only summary statistics were meta-analysed in METAL under the STDERR scheme, weighting effect estimates by the inverse of their standard errors. Between-sex heterogeneity was assessed for each variant using Cochran’s Q statistic.

We defined sex-differential loci as variants reaching genome-wide significance in at least one sex (p<5×10^−8^) with significant between-sex heterogeneity (phet<5×10^−8^). Independent regions were defined as ±500-kb windows around each heterogeneity-significant variant, with overlapping windows merged. The MHC region was treated as a single region. Within each region, the variant with the smallest heterogeneity p-value was designated the sentinel. For each sentinel, sex-specific effect estimates were extracted and classified as male-only, female-only, or significant in both sexes, and variants with opposite effect directions between sexes were additionally flagged. Finally, sex-differential sentinels were cross-referenced against credible sets from the sex-combined GWAS to determine which sex-differential loci were also detected in the combined analysis.

### Mapping of druggable loci

To systematically test whether embeddings associated with genomic loci mimicking drug action, we compiled a list of 426 loci proxying the effect of 724 approved or clinically advanced (reached phase 3) drugs across 106 indications. To this end, we parsed results collated in the GWAS Catalog (download: 22/05/2025) against drugs and their targets reported in the OpenTargets platform^46^ and retained only locus – indication combinations with strong evidence that: 1) the locus encodes for a canonical drug target, a ligand – receptor pair of the canonical target, or complex partner of the canonical target, 2) is associated with the indication of the respective drug, and 3) the variant likely acts through any of the targets listed under 1). Across each of the 426 loci, we performed systematic colocalization testing as implemented in the R package ‘coloc’ (v5.2.3)^55^ across all embeddings. We used default priors, apart from p12 that we adopted to 5×10^−6^ following previous work^56^. Colocalisation was only performed if any SNP in the region associated with the embedding with p<10^−5^ to minimize computational burden. We subsequently filtered colocalization results at PP≥80% for a shared genetic signal between the embedding and drug indication and further ensured that respective regional sentinel variants were in high LD (r^2^>0.8) to flag potentially artificial results. We applied the identical pipeline to GWAS summary statistics for ICD-10 based diagnoses published previously^24^.

### Cox proportional-hazards models for disease onset

We collated a comprehensive phenotypic dataset from the UK Biobank baseline visit, described in detail previously^35^. This dataset contained genetically inferred ancestral group, diagnoses recorded up to the baseline visit (n=1,197), self-reported medications (n=695), clinical-chemistry biomarkers (n=28), blood-cell counts (n=29), measures of body composition (n=20), bone (n=7) and cardiovascular (n=3) health, diet (n=23), socioeconomic (n=9) and general-health (n=4) status, basic demographics (n=5), pulmonary function (n=8) and operational parameters (n=5). Apart from medications and diseases, each variable was used as an exposure in Cox proportional- hazards models testing its association with the first reported onset of 636 ICD-10-coded diagnoses. Continuous exposures were rank-inverse-normal transformed. Models were adjusted for age and sex and stratified by recruitment site, and we excluded participants with a diagnosis within six months of baseline to mitigate reverse causation. For each exposure we quantified the increment in Harrell’s C-index over the adjustment set (ΔC-index), where a disease’s Delphi-2M baseline linear predictor was available, we additionally computed the increment over a model already containing that predictor and the adjustment set (the marker’s gain beyond Delphi-2M) and the gain of the Delphi-2M predictor itself over the adjustment set. Baseline C-indices were stored per sex-subset and so used all participants of that subset, whereas the full models used each exposure’s complete cases, the resulting ΔC-index is therefore a close but not exactly matched estimate. This ΔC-index was used to select exposures that independently explained variation in Delphi-2M’s embedding-derived AUCs by statistical fine-mapping, using a set-up analogous to the genetic analysis, supplemented with features from a partial-correlation network additionally adjusted for age and sex across the 636 diagnoses over the lifespan. A marker’s pleiotropy was defined from the Cox models as the number of distinct diagnoses with which it was associated at a Bonferroni-corrected threshold (p<3.3×10^−6^) and its primary disease as the association with the largest C-index increment. To quantify how much of each exposure the representation already encodes, we regressed every exposure on the 120 embeddings by linear regression and recorded the explained variance (R²) separately by sex, pooling across sexes for reporting only.

### Contesting the predictive capacity of Delphi-2M

To test whether biomarkers carried predictive value over and above Delphi-2M, we used each individual’s per-disease baseline log-rate from the model as a fixed linear predictor (the Delphi linear predictor) in the held-out evaluation set (continuous biomarkers, n=92,643, proteins, the UK Biobank Olink subcohort, n=7596). Because the C-index over the full follow-up forces the Delphi predictor into a single proportional-hazards term, we confirmed incremental value with a horizon-specific delta-AUC. Individuals were classified as cases (incident diagnosis within a 10- year horizon) or controls (event-free with at least the horizon of follow-up), excluding prevalent diagnoses, events within the first six months, and participants censored before the horizon. Within 5-year-age × sex strata (strata defined on sex for sex-combined analyses) we compared the discrimination of the Delphi linear predictor alone with that of a combined score, the combined score (a logistic model of incident status on the Delphi linear predictor and the biomarker) and the component AUCs were estimated out of sample by 5-fold cross-fitting, the Delphi-only baseline being the raw linear predictor. Per-stratum AUC differences were obtained by DeLong’s method for paired AUCs^57^ and aggregated by case-weighting. We separately recorded each biomarker’s own gain over an age-and-sex baseline using the same cross-fitted procedure, with the exposure score oriented out-of-fold so that protective markers are not scored as negative discrimination. We deliberately did not construct an extended lifestyle baseline. Body-mass index, alcohol consumption and smoking are themselves input tokens to Delphi-2M and are therefore represented within its linear predictor by construction, a baseline incorporating them would have been a sub-model of the predictor under evaluation, entangling “gain over baseline” with “gain over Delphi-2M”, and would in any case leave unchanged the marker-versus-Delphi contrast on which our conclusions rest, which does not depend on the demographic baseline. Because these lifestyle tokens were supplied to Delphi-2M with deliberately randomised timing^6^, they were represented in degraded form, the age-and-sex baseline used for the secondary own-gain anchor is correspondingly not adjusted for adiposity, so this anchor may have overstated the standalone value of the minority of biomarkers that are largely body-composition readouts, without affecting the primary contrast.

From the collated Cox, delta-AUC and reconstruction outputs we derived three quantities per feature, jointly across clinical biomarkers and proteins. First, reconstruction: the variance of each exposure explained by the 120 embeddings (R^2^, within sex), summarised as the median, interquartile range and the proportion of features reconstructed above fixed R^2^ thresholds. Second, absorption: across all robustly estimated marker–disease pairs, the median incremental discrimination over Delphi-2M (ΔAUC) relative to the median gain over an age-and-sex baseline, expressed as the proportion of a marker’s demographic gain absorbed by the model, together with the rank correlation between reconstruction and both quantities. Third, residual specificity: for each marker, the residual ΔAUC over Delphi-2M at the single disease it most strongly defined, where that disease was selected by the marker’s Cox C-index increment and deliberately not by its ΔAUC, to avoid selection on the discrimination outcome. Marker - disease pairs were retained only where the incident-case count was ≥100. Complementarily, for each disease we recorded the largest ΔAUC over Delphi-2M achieved by any single marker (the optimistic single-marker ceiling, estimated out of sample per pair) and classified the winning marker as a diagnostic analyte, an organ-function measure, or a secreted/tissue-leakage protein. Finally, to test whether the model absorbs proportionally more of a marker’s value the more diseases it was associated with, we related each marker’s pleiotropy (the number of distinct diseases for which it reached a Cox association at a Bonferroni threshold) to the value absorbed by Delphi-2M (the gain over demographics minus the residual over Delphi-2M). We note that pleiotropy, reconstruction, and residual value are mutually positively correlated, so this relationship should be considered descriptive rather than as an independent explanatory axis.

