## Supplementary figures for "Genetic decoding reveals druggable biology implicitly learned by a medical-history foundation model"

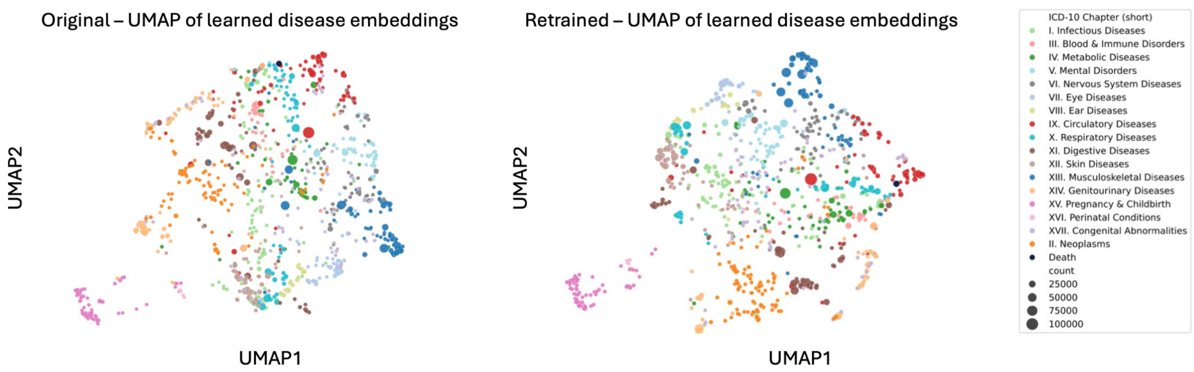


**Supplementary Figure 1 UMAP projection of token embeddings, coloured by disease chapter.** The token (disease) embeddings derived from the retrained Delphi-2M (right) showed a distribution similar to that reported in the original study (left), both in terms of within-chapter and between chapter distance.

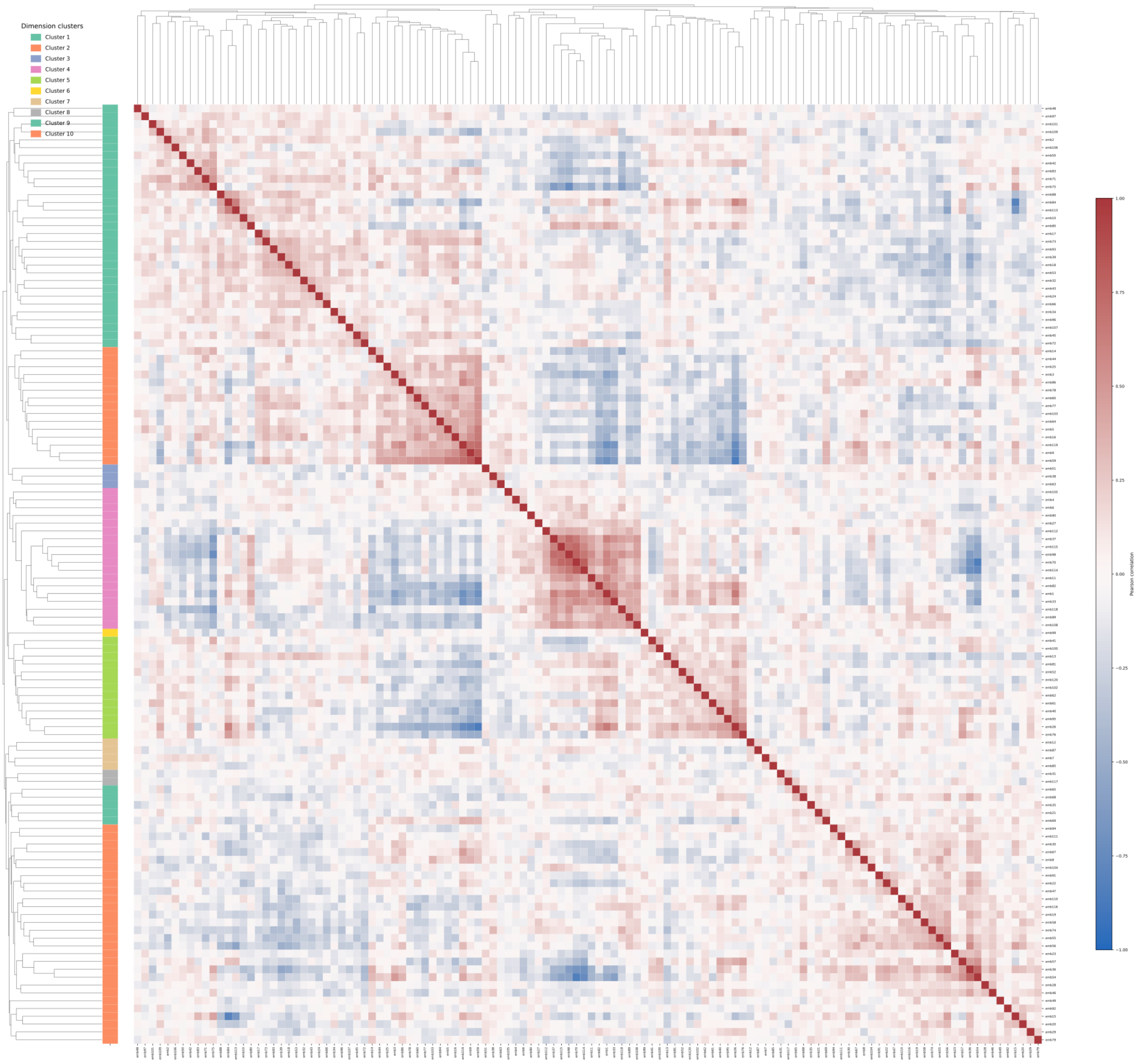

**Supplementary Figure 2 Correlation structure among 120 patient embedding dimensions.** Heatmap showing pairwise Pearson correlation coefficients between 120 embedding dimensions across individuals. Dimensions were grouped by hierarchical clustering using average linkage, resulting in 10 clusters indicated by the coloured annotation.

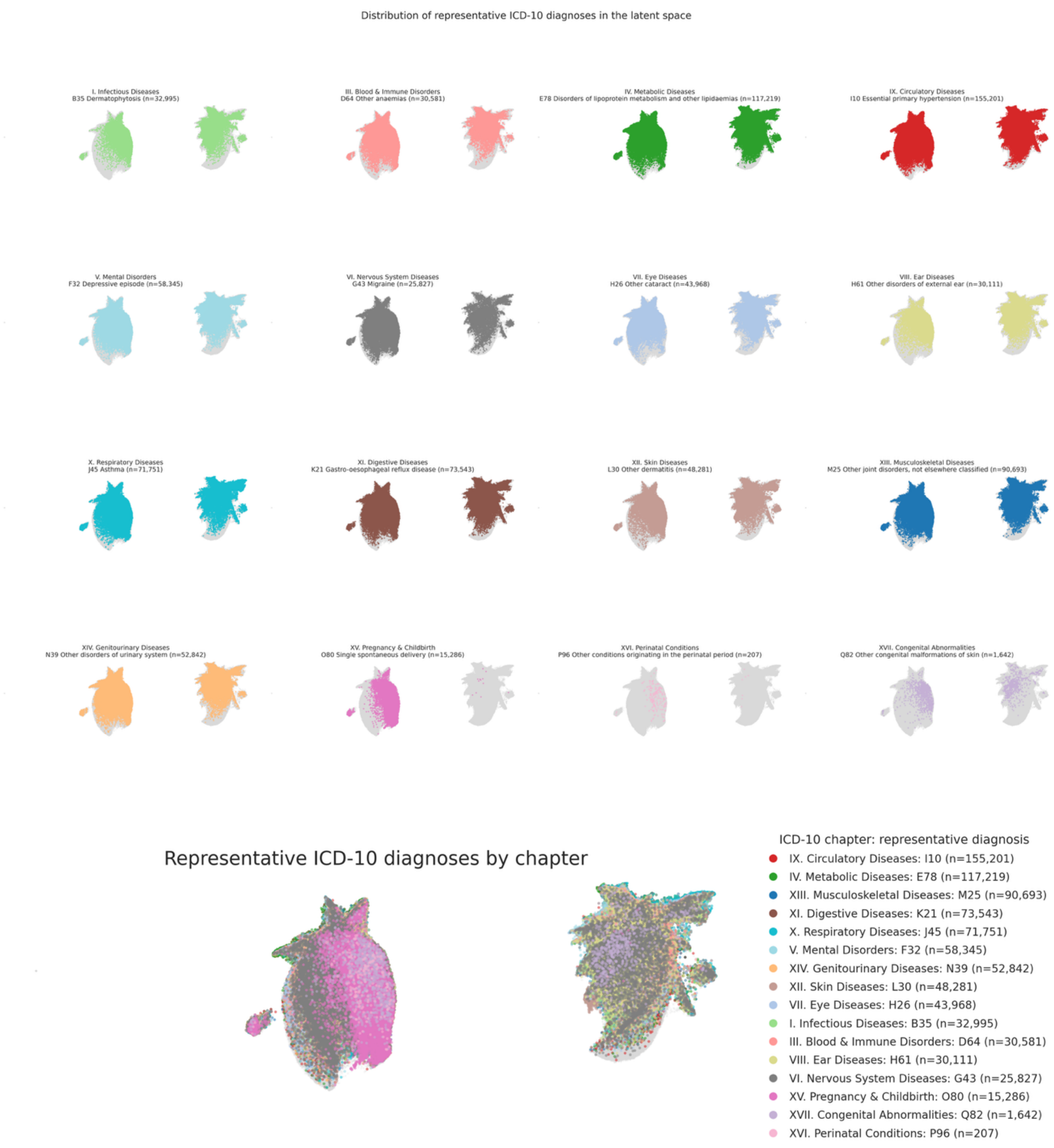


**Supplementary Figure 3 Lack of diagnosis-separation in patient embedding space** UMAP projection of patient embeddings, coloured by representative ICD-10 diagnoses. For each ICD-10 chapter, the diagnosis recorded in the largest number of individuals was selected. Diagnoses are shown in separate panels, with coloured by chapter and all other individuals shown in grey (upper). The same representative diagnoses are displayed in a single UMAP, coloured by chapters (lower).
